# Diabetic macular thickness diagnostic guideded by optical coherence tomography using artificial intelligence methods: a scoping review protocol

**DOI:** 10.1101/2020.10.17.20214288

**Authors:** Fabiana Brasileiro, Frederico M. Bublitz

## Abstract

**Introduction:** diabetic retinopathy remains the mainly cause of blindness worldwide and its diagnoses and assessment is poorer as the countryside the patient is. To minimize de boundaries of health care and improve the accuracy how the treatment is stablished, the artificial intelligence (AI) techniques have been studied years ago using fundus phothographies for instance. This scoping review aims to study these AI techniques applied in Opctical Coherence Tomography (OCT) images.

**Methods:** This scoping review will follow the methodology framework defined in “Scoping studies: advancing the methodology”. In this methodological framework, six stages are proposed for scoping review

**Studies:** identifying the research question; identifying relevant studies; study selection; charting the data; collating, summarizing, and reporting the results; and consultation. The research questions aim to investigate what are the methods, and techniques used in artificial intelligence due to perfmorm a clinical diagnostic using OCT scanners. The team will focus on the Scopus Document Search e PubMed (Medline). The search query is a combination of terms related to Diabetic Retinopathy AND Artificial Intelligence.

**Ethics and Dissemination:** This is a scoping review study and there is no requirement for ethical approval, as primary data will not be collected. The results from this scoping review will be published in a peer reviewed journal and reported at scientific meetings. We intend to share the results with the Ophthalmologists.

**ARTICLE SUMMARY:** *Strengths and Limitations of this study:* This protocol uses a comprehensive approach, relating one research question, that will enable the identification of the main gaps and opportunities in the area;

- Due to the comprehensiveness of this review protocol, it can serve as the basis for future works, with more specific scope, and the proposal of standards;
- The investigation of who are the mainly artificial intelligence skills will allow other researchs in this area;
- The inclusion of aspects of data governance in the protocol will serve as the basis for establishing an assessment model of artificial intelligence and diabetic macular thickness;
- One weakness of this review protocol was the number of the studies including glaucoma in their criterias.

## INTRODUCTION

Diabetic retinopathy is still one of the manly cause of irreversible blindness in the world[1] and macular edema leads the visual impairment. To prevent blindness the screening and early treatment are the best aproach[2].

The gold standard macular thickness assessment is provided by OCT images. Artificial intelligence may improve its accuracy.

## METHODS AND ANALYSIS

This scoping review will follow the methodology framework defined by Levac et al.[3]. In this methodological framework, six stages are proposed for scoping review studies: 1) identifying the research question; 2) identifying relevant studies; 3) study selection; 4) charting the data; 5) collating, summarizing, and reporting the results; and 6) consultation (optional).

Following this section, the stages of the methodological framework for conducting a scoping study are presented.

### Identifying the research question

The aim of this study is to identify what are the thchniques of artificial intelligence used to allow computers systems to identify diabetic retinopathy as well its complication called diabetic macular thickness that cause visual impairment.

### Identifying relevant studies

This review will be conducted entirely using electronic databases. The following databases were used: Scopus Document Search and PubMed (Medline)^1^. These electronic databases were chosen because they cover a vast amount of areas with relevant works related to the scope of this review, such as technology and health. One important note about the choice of the data sources is that the Springer was initially considered to be included; however, most of the papers returned were not relevant as it was not possible to perform an advanced search. In addition to that, the papers returned by Scopus already contemplate papers from Springer.

The researcher of this scoping review has large experience in ophthalmology specially in diabetic retinopathy. For this review, she has already performed some rounds for tuning the search string. The main terms related to artificial intelligence and diabetic retinopathy. The terms used during this round were: “diabetic retinopathy”, “diabetic macular thickness”, “artificial intelligence”. (there’s no relevant plural ways).

After two rounds of using different combination involving these previous mentioned, and other related, terms and evaluating the relevance of some of the returned papers. Actually, we get 5 papers as control papers, these control papers were articles that we read during these rounds and pre-classified them as relevant, and that should appear in the search. The final search query, used in the Scopus Document Search database as an example, is TITLE-ABS-KEY (“Optical Coherence Tomography” OR “OCT” AND (“Diabetic Retinopathy” OR “Diabetic Macular Thickness” AND “Artificial Intelligence”.

### Study selection

The focus of this review is to have a comprehensive view on what artificial intelligence methods have been used to perform retinal diagnosis in diabetic macular edema.

The team met to discuss the inclusion and exclusion criteria. From these meetings the team has decided that the main restriction with regard to domain is that the paper must delivery the diabetic macular edema enrolled and some artificial intelligence method used. The other restrictions are more generic and take into account criteria like availability, language, and the type of publication (i.e., primary or secondary study). The following inclusion (INC) and exclusion (EXC) criteria were defined:

- INC.01. - Domain: The paper addresses an artificial intelligence method enrolled in the diabetic macular edema assessment using OCT images;
- INC.02. - Language: The paper must be written in English;
- INC.03. - Time: There are no time restrictions. This option differs a little bit from traditional scoping reviews, which typically include papers from the last five years, but the team decided to include any solution despite the date of publication to perceive how the theme has evolved over time;
- INC.04. - Study Design: The paper presents a primary study. The team is looking for primary research papers. The team already performed a screening on secondary studies, and, although there are good reviews, there are still gaps that the team overcame by analyzing primary studies;
- INC.05. - Availability: The full paper is available. Although the team has access to a considerable amount of datasets, some search engines (e.g. Scopus Document Search) point to a third data source where some of the studies may not be available (or only accessible under purchase);
- EXC.01. - Duplicated: The team is using a set of datasets for searching, which may increase the possibility of the paper appearing more than one time. In this case, the team will eliminate the duplicates;
- EXC.02. - not matching all the inclusion criteria: Any paper that does not match all the inclusion criteria will be excluded from the review. The only exception is regarding the domain inclusion criteria, in this case, the paper must fit at least one of them (if not all).
- EXC.03. -were excluded too, studies that present the therms: glaucoma, optical nerve head and age related macular degeneration.

Given the presented inclusion and exclusion criteria, the team will start the screening and eligibility phases based on the flow diagram from PRISMA Statement [4]. For these phases, each paper will be read by two members of the team, and in case of a conflicting decision another member will read the conflicting paper and discuss it with the other members to solve the conflict. Hence, the team will adhere to the following steps for the review: i) looking for duplicates, maintaining only one version of them; ii) performing a fast reading on abstract, introduction and conclusion and exclude those studies that clearly fall on one of the exclusion criteria; and iii) the team will read the paper and exclude those studies that fall on one of the exclusion criteria, explaining the reasons for exclusion.

Regarding the quality assessment of the publication source and its impact, despite that in a first moment, this information will not be used to exclude papers, it is very important to minimize some bias that publications could bring to the review.

### Charting the data

This stage consists of mapping the information that will be extracted from the primary studies being analyzed[5]. For this stage, the team is using a data chart form using the record information adapted from Crick et al. work[6], plus information related to the scope of this review, as follows:

- Bibliographical information: title of the article, authors(s), country, year, as well as the quality of the publication source and its impact;
- Study information: aims of the study, methodology, outcome measures (if applicable), important results;
- AI information methods;
- Team considerations: strengths of the study, limitations of the study, and conclusion.

For extracting the information, each paper will be read by two members of the team; each member will collect the data independently. After that, the collected data for the two distinct members will be merged, and for the conflicting cases, a third member will be consulted to diminish the conflict.

### Collating, summarizing, and reporting the results

Following the recommendations provided by Levac et al. [3] this stage will be broken into three steps, as follows:

- Analysis: for this step, the team will provide qualitative analysis that will use the data from papers and team considerations to extract information that can give a better overview of subjective concepts and opens up new opportunities of studies, for example, we aim to have a deep understanding of artificial intelligence methods;
- Reporting: for this step the team intend to present a table of strengths and gaps in the evidence; moreover, the structured results combined with the analysis will serve as basis for the creation of a paper;
- Implications for future research, practice and policy: we aim to discuss these results with the ophthalmologists community.

### Patient and Public Involvement

Patients and or public were not involved.

## DISCUSSION

This review will alow to identify the best AI techniques used in diabetic retinopathy diagnosis from OCT images. It will allow to realize the gaps in this area.

## Data Availability

https://www.ncbi.nlm.nih.gov/pubmed

## AUTHOR CONTRIBUTIONS

For this article we had the following author contributions: conceptualization, methodology, and writing--original draft preparation, Frederico M. Bublitz and Fabiana Brasileiro; writing--review and editing Fabiana Brasileiro.

## ETHICS AND DISSEMINATION

This is a scoping review study and there is no requirement for ethical approval, as primary data will not be collected. The results from this scoping review will be published in a peer-reviewed journal and reported at scientific meetings.

## Footnotes

1 https://www.ncbi.nlm.nih.gov/pubmed/

